# Association of polygenic risk scores with low-density lipoprotein cholesterol levels and control: findings from the Hispanic Community Health Study/Study of Latinos (HCHS/SOL)

**DOI:** 10.64898/2026.09.16.26363269

**Authors:** Christina G Hutten, Tamar Sofer, Brian Spitzer, Molly Scannell Bryan, Jiehuan Sun, Victoria W Persky, Martha L Daviglus, Maria Argos

## Abstract

**Background:** Hypercholesterolemia is a prevalent cardiovascular (CVD) risk factor among Hispanics/Latinos; however, rates of cholesterol awareness and treatment are low in this population. Using a polygenic risk score (PRS) associated with low-density lipoprotein cholesterol (LDL) levels may help identify those who would benefit most from statin therapy. Previous LDL-PRS have been developed but they have not been properly evaluated for use in a highly diverse Hispanic/Latino population.

**Methods:** The Hispanic Community Health Study/Study of Latinos (HCHS/SOL) is a prospective study that enrolled 16,415 Hispanic/Latino adults aged 18-74 years from four U.S. communities in 2008-2011. We assessed the association of twenty PRS with baseline LDL levels and LDL control among 11,669 HCHS/SOL participants with complete data and consent to conduct genetic research. Weighted PRS were calculated using effect estimates obtained from the PGS Catalog. Multivariable linear regression analysis was used to derive effect estimates (betas (β), 95% confidence intervals (CI)) for the association between each PRS, modeled continuously and by quintiles, with baseline LDL levels. Multivariable logistic regression was used to derive odds ratios and 95% CI for the association between each PRS and LDL control in statin users. Models were adjusted for predefined confounders and accounted for survey weights.

**Results:** A PRS developed with PolyFun-pred, using European GWAS summary statistics, provided the largest incremental improvement for predicting LDL levels for the full sample (ΔR^2^ 0.114), Caribbean background (ΔR^2^ 0.096), majority African ancestry (ΔR^2^ 0.098), and majority Amerindian ancestry subgroups (ΔR^2^ 0.123). PRSCSx-EUR, using multiple ancestry GWAS and weights, improved PRS performance most for the Mainland background (ΔR^2^ 0.090) and majority European ancestry (ΔR^2^ 0.061) subgroups. For every one standard deviation increase across the 20 PRS asssesed, LDL increase varied between 3 and 14mg/dL. Among 1,323 statin users, the odds of LDL control were greatly reduced with increasing PRS but differed between PRS and Hispanic/Latino background groups and genetic ancestry groups.

**Conclusion:** A polygenic risk score prioritizing functional annotations demonstrated superior performance for predicting LDL levels and LDL control across Hispanic/Latino subgroups. These results emphasize that further development of PRS is needed for improved risk prediction before clinical application in diverse populations.

**Clinical Perspective:** *What is new?:*

- Twenty previously validated polygenic risk scores associated with low-density lipoprotein (LDL) were found to be positively associated with LDL in a diverse population of Hispanics/Latinos living in the U.S. who have been historically underrepresented in genomic research.
- PRS developed using Hispanic/Latino GWAS summary statistics did not improve performance in all Hispanic/Latino subgroups. The PolyFun-pred PRS developed using European GWAS summary statistics and prioritizing functional annotations, improved performance most for subgroups historically underrepresented in genomic research.
- The higher the PolyFun-pred PRS, the lower the likelihood participants had control of LDL levels while taking statins for both Caribbean and Mainland subgroups.
- What are the clinical implications? (<100 words)
- The PolyFun-pred PRS shows promise to identify those at higher risk for increased LDL levels and may benefit from statin therapy; however, additional management may be necessary for those with high PolyFun-pred PRS because statin therapy was less likely to bring LDL under control.
- The Hispanic/Latino population is genetically heterogeneous and should not have a one-size-fits-all approach when it comes to precision medicine.
- To be clinically useful, further research is needed to identify a PRS that will provide a diverse population of Hispanics/Latinos with the same benefit from pharmacogenomics and precision medicine as European populations to prevent widening health disparities.

## Introduction

In the Hispanic Community Health Study/Study of Latinos (HCHS/SOL), hypercholesterolemia is the most prevalent cardiovascular risk factor among men and the second most prevalent risk factor among women (Daviglus et al., 2012, 2014). Despite the high burden in this population, rates of high cholesterol awareness and treatment are low in this population (Rodriguez et al., 2015). In HCHS/SOL, about half of the participants were aware of having high cholesterol, less than a third of those were on treatment, and only about one-third had adequately controlled cholesterol levels (Qureshi et al., 2017; Rodriguez et al., 2015). In a separate study, Hispanics/Latinos over 40 years of age with a history of atherosclerotic cardiovascular disease had 31% lower odds of current statin use than non-Hispanic whites (Ngo-Metzger et al., 2019).

Elevated low-density lipoprotein cholesterol (LDL) is a modifiable risk factor for cardiovascular disease (CVD) and stroke (Benjamin et al., 2019; Liu et al., 2017) and treatment with statins can reduce the risk of cardiovascular events (Postmus et al., 2014; Wei et al., 2018). Statin therapy (hydroxymethyl-3-methylglutaryl coenzyme A reductase inhibitors) can reduce LDL levels by 30-55%, the risk of major coronary events by 27%, and all-cause mortality by 15% (Baigent et al., 2005; Baigent et al., 2010; Ciuculete et al., 2017; Leusink et al., 2016; Mihaylova et al., 2012; Postmus et al., 2014; Taylor et al., 2013). The use of statins is highly prevalent worldwide (Laleman et al., 2018); however, an individual’s response to therapy varies due to both genetic and environmental factors (Chasman et al., 2012; Kim et al., 2020; Leusink et al., 2016; Mangravite et al., 2006; Smit et al., 2016; Wei et al., 2018). Reduction in LDL in response to statin therapy has been shown to vary across racial/ethnic groups (Oni-Orisan et al., 2018; Simon et al., 2006).

Polygenic risk scores (PRS) constructed using hundreds to millions of single nucleotide polymorphisms (SNPs) associated with LDL levels have been compiled in the PGS Catalog. PRS, developed using effect estimates from various genome-wide association studies (GWASs) with differing statistical methods, aimed to increase portability across diverse ancestral populations (Graham et al., 2021; Klarin et al., 2018; Ma et al., 2022; Privé et al., 2022; Shim et al., 2023; Trinder et al., 2020; Weissbrod et al., 2022). Few LDL PRS have been assessed in the Hispanic/Latino population, particularly Hispanics/Latinos with diverse ancestral backgrounds. This analysis evaluated 20 PRS from the PGS Catalog, using differing numbers of SNPs, ancestral GWAS backgrounds, and statistical methods to identify which best associated with baseline LDL levels and LDL control among statin users in the diverse HCHS/SOL cohort. We sought to determine whether Hispanic/Latino individuals identified to be at the highest risk of elevated LDL levels according to this PRS could also benefit most from statin treatment.

## Methods

### Study population and data collection

Existing longitudinal data collected as part of the Hispanic Community Health Study/Study of Latinos (HCHS/SOL) cohort and existing genotype data were utilized in this analysis. The HCHS/SOL is a multicenter longitudinal study to evaluate and identify risk and protective factors in relation to the health of U.S. Hispanics/Latinos (LaVange et al., 2010; Sorlie et al., 2010). It is one of the largest studies of Hispanic/Latino health in the U.S., with 16,415 participants aged 18-74 years enrolled through four field centers. Recruitment was conducted using a two-stage area probability sampling of households in Chicago, San Diego, Bronx, and Miami (LaVange et al., 2010; Sorlie et al., 2010). Institutional Review Board (IRB) approval was obtained at each center’s respective IRB, and all participants provided written informed consent in the language of their preference (English or Spanish). Of the 16,415 HCHS/SOL participants, approximately 77% consented to the use of their DNA samples in genetic research by HCHS/SOL affiliated investigators. Participants taking lipid-lowering medications other than statins were excluded (n = 128). The analytic sample consisted of participants with complete baseline data on LDL cholesterol, statin use, and key covariates (n=11,669) (**Supplemental Figure 1**).

### Clinical evaluations in the HCHS/SOL

Participants underwent an extensive clinical exam and assessments at baseline (Visit 1: 2008-2011). Exams were conducted by trained/certified technicians and health interviewers at each field center according to standard protocols (HCHS/SOL manuals). Participants were asked to fast and abstain from smoking 12 hours prior and avoid vigorous physical activity on the morning of the examination. Anthropometric characteristics were measured, including body weight to the nearest 0.1 kg and height to the nearest centimeter. Body mass index (BMI) was calculated as weight in kilograms divided by height in meters squared. Three seated blood pressure measurements were obtained after a 5-minute rest using an oscillometric automated sphygmomanometer. The average of the second and third measurements was calculated for use in analyses. Diabetes was defined using American Diabetes Association guideline lab criteria (American Diabetes Association, 2010), taking into account plasma glucose levels adjusted for fasting time, post-OGTT glucose levels, Hemoglobin A1c (HbA1c) percentages, and self-report of diabetes.

### Laboratory evaluation in the HCHS/SOL

Fasting blood samples collected from the participants were shipped to the HCHS/SOL Central Laboratory at the University of Minnesota, where the following parameters were measured: total cholesterol using a cholesterol oxidase enzymatic method; high-density lipoprotein (HDL) cholesterol using a direct magnesium/dextran sulfate method; plasma glucose using a hexokinase enzymatic method; serum triglycerides using a glycerol blanking enzymatic method (Roche Diagnostics, Indianapolis, IN); low-density lipoprotein (LDL) cholesterol was calculated using the Friedewald equation (Friedewald et al., 1972); HbA1c was measured using a Tosoh G7 Automated HPLC Analyzer (Tosoh Bioscience) (Daviglus et al., 2012).

### Covariates

Information collected via an interviewer-administered questionnaire at Visit 1 included: age, sex, Hispanic/Latino background (Cuban, Dominican, Mexican, Puerto Rican, Central American, South American, and other), field center, alcohol use, and smoking history.

### Outcome

Among the 16,415 HCHS/SOL Visit 1 participants, low-density lipoprotein (LDL) cholesterol levels (mg/dL) were available for 12,029 (**Supplemental Figure 1**). Statin therapy was verified from information scanned and transcribed by study personnel from medications that were taken for the 4 weeks prior, brought in by participants to their Visit 1 appointment. For participants with verified statin use at enrollment, baseline LDL levels were multiplied by 1.43 to account for a 30% reduction of LDL associated with statin use (Trinder et al., 2020). LDL control was assessed among those taking statins (n=1323) at Visit 1. LDL control was defined as observed LDL levels <130 mg/dL (Stone et al., 2014) or LDL < 100mg/dL for those with diabetes (American Diabetes Association, 2011; Casagrande et al., 2017). Sensitivity analysis was conducted to assess for modification of the association between the PRS and LDL control by diabetes status among statin users using control as defined above and updated guidelines defined as <100mg/dL and <70 mg/dL for those with diabetes (Grundy et al., 2019).

### Genotyping, Imputation and Quality Control

HCHS/SOL individuals who consented to participation in genetic studies at Visit 1 had DNA extracted from whole blood samples and genotyped using a customized HCHS/SOL Illumina Omni 2.5 M array (HumanOmni2.5-8 v.1-1) (Dunn et al., 2018; Rosenberg et al., 2003; Sofer et al., 2019). Standard quality assurance and quality control measures were applied to generate recommended variant- and sample-level quality filters (Laurie et al., 2010; Sofer et al., 2019). There were 2,232,944 genetic variants that passed quality filters and were informative that proceeded for imputation (Conomos et al., 2016). Imputation was conducted via the University of Michigan TOPMed 2.0 imputation server (Das et al., 2016; Taliun et al., 2021). Imputation quality (R^2^) was reported for each variant. Proportions of genetic ancestry were calculated using ADMIXTURE software (Alexander et al., 2009; Conomos et al., 2016).

### Polygenic Risk Score

PRSs associated with LDL cholesterol measurement (EFO_0004611) were selected from the PGS Catalog (Lambert et al., 2021) in order to assess the performance of various methods of PRS construction with varying numbers of SNPs included and differing GWAS ancestry populations (**Table 1**). Only autosomal variants with MAF ≥ 0.01 and imputation quality R^2^ ≥ 0.8 were used for PRS construction. Variant weight files were downloaded from the PGS Catolog (Lambert et al., 2021). Weights were lifted over from hg19 to hg38 in order to match the target data when necessary. Multi-allelic variants were discarded.

**Table 1.**
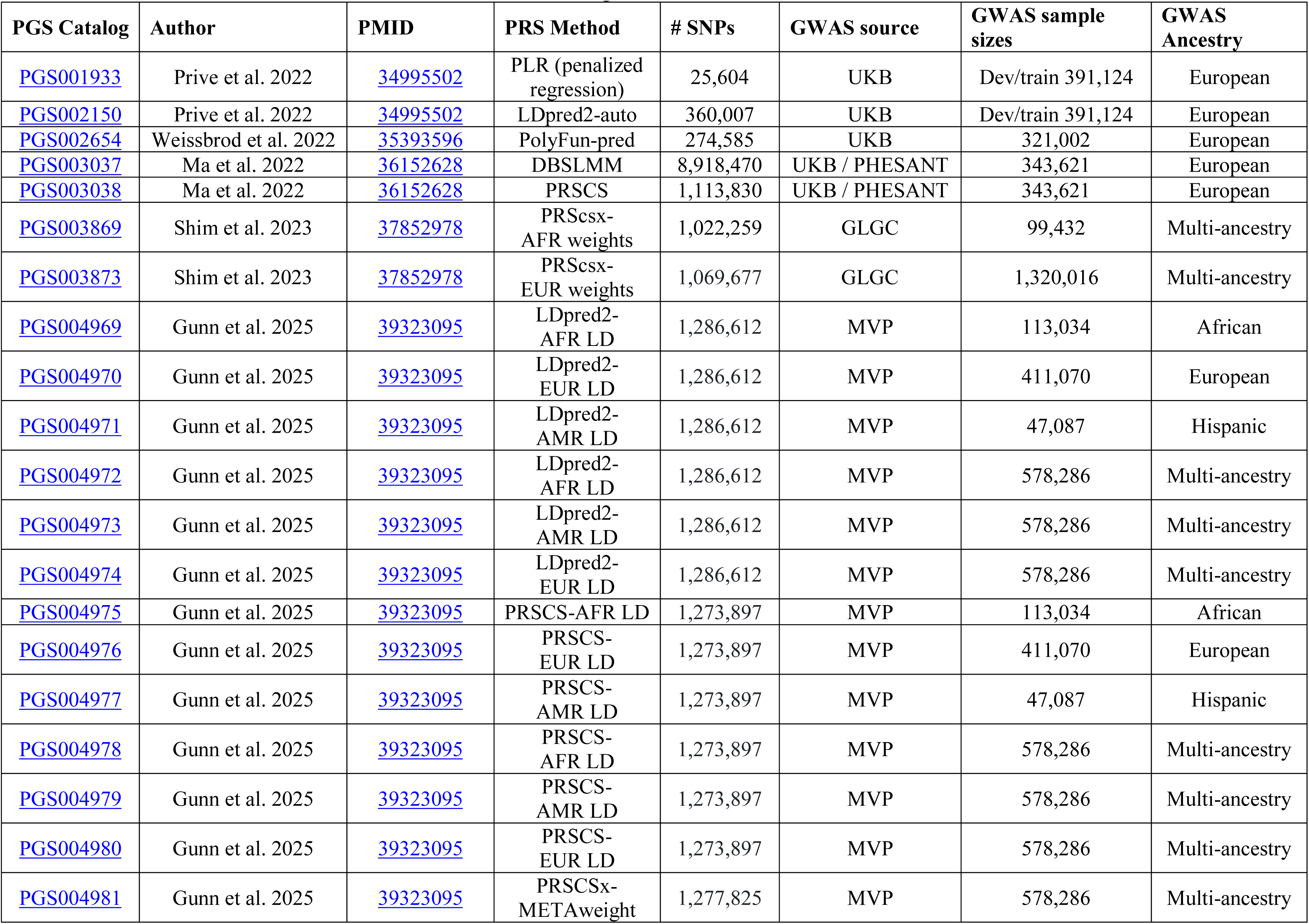
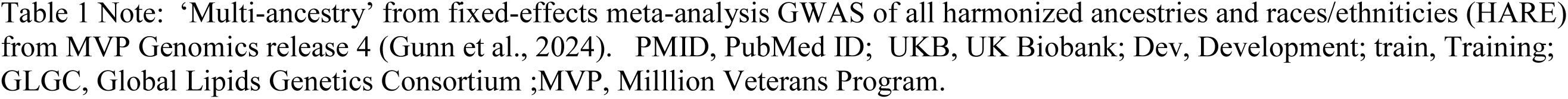
Characteristics of PRS selected from the PGS Catalog.

Note: ‘Multi-ancestry’ from fixed-effects meta-analysis GWAS of all harmonized ancestries and races/ethnicities (HARE) 905 from MVP Genomics release 4 (Gunn et al., 2024).
| PGS Catalog | Author | PMID | PRS Method | # SNPs | GWAS source | GWAS sample sizes | GWAS Ancestry |
| --- | --- | --- | --- | --- | --- | --- | --- |
| PGS001933 | Prive et al. 2022 | 34995502 | PLR (penalized regression) | 25,604 | UKB | Dev/train 391,124 | European |
| PGS002150 | Prive et al. 2022 | 34995502 | LDpred2-auto | 360,007 | UKB | Dev/train 391,124 | European |
| PGS002654 | Weissbrod et al. 2022 | 35393596 | PolyFun-pred | 274,585 | UKB | 321,002 | European |
| PGS003037 | Ma et al. 2022 | 36152628 | DBSLMM | 8,918,470 | UKB / PHESANT | 343,621 | European |
| PGS003038 | Ma et al. 2022 | 36152628 | PRSCS | 1,113,830 | UKB / PHESANT | 343,621 | European |
| PGS003869 | Shim et al. 2023 | 37852978 | PRScsx-AFR weights | 1,022,259 | GLGC | 99,432 | Multi-ancestry |
| PGS003873 | Shim et al. 2023 | 37852978 | PRScsx-EUR weights | 1,069,677 | GLGC | 1,320,016 | Multi-ancestry |
| PGS004969 | Gunn et al. 2025 | 39323095 | LDpred2-AFR LD | 1,286,612 | MVP | 113,034 | African |
| PGS004970 | Gunn et al. 2025 | 39323095 | LDpred2-EUR LD | 1,286,612 | MVP | 411,070 | European |
| PGS004971 | Gunn et al. 2025 | 39323095 | LDpred2-AMR LD | 1,286,612 | MVP | 47,087 | Hispanic |
| PGS004972 | Gunn et al. 2025 | 39323095 | LDpred2-AFR LD | 1,286,612 | MVP | 578,286 | Multi-ancestry |
| PGS004973 | Gunn et al. 2025 | 39323095 | LDpred2-AMR LD | 1,286,612 | MVP | 578,286 | Multi-ancestry |
| PGS004974 | Gunn et al. 2025 | 39323095 | LDpred2-EUR LD | 1,286,612 | MVP | 578,286 | Multi-ancestry |
| PGS004975 | Gunn et al. 2025 | 39323095 | PRSCS-AFR LD | 1,273,897 | MVP | 113,034 | African |
| PGS004976 | Gunn et al. 2025 | 39323095 | PRSCS-EUR LD | 1,273,897 | MVP | 411,070 | European |
| PGS004977 | Gunn et al. 2025 | 39323095 | PRSCS-AMR LD | 1,273,897 | MVP | 47,087 | Hispanic |
| PGS004978 | Gunn et al. 2025 | 39323095 | PRSCS-AFR LD | 1,273,897 | MVP | 578,286 | Multi-ancestry |
| PGS004979 | Gunn et al. 2025 | 39323095 | PRSCS-AMR LD | 1,273,897 | MVP | 578,286 | Multi-ancestry |
| PGS004980 | Gunn et al. 2025 | 39323095 | PRSCS-EUR LD | 1,273,897 | MVP | 578,286 | Multi-ancestry |
| PGS004981 | Gunn et al. 2025 | 39323095 | PRSCSx-METAweight | 1,277,825 | MVP | 578,286 | Multi-ancestry |

Scores were obtained for each individual using PLINK 1.9 function ‘--score’ then summed over the autosomes. Standardized PRSs were calculated such that the PRSs had mean zero and standard deviation 1 for the full HCHS/SOL sample with genetic data and non-missing PRS sums (n = 12,313). Sensitivity analysis standardizing PRS by subgroup (Mainland and Caribbean) was performed for all PRS. The 20 PRSs are summarized in **Table 1** and the methodologies used for construction are summarized below:

1. PRS-CS (Ge et al., 2019) [PGS003038 (Ma et al., 2022), PGS004975, PGS004976, PGS004977, PGS004978, PGS004979, PGS004980, PGS004981 (Gunn et al., 2025)]: Estimates posterior SNP effect sizes under a continuous shrinkage prior, integrating summary statistics with an external linkage disequilibrium (LD) reference panel to account for correlations between variants (Ge et al., 2019).
2. Penalized Regression (PRL) [PGS001933 (Privé et al., 2022)]: Using ‘bigstatsr’ R package, PRS were computed with lasso-penalized regression, modeling phenotype as a function of genotypes and covariates while shrinking SNP effect sizes to prevent overfitting. Models allowed variant-specific penalties (e.g. scaling by SNP variance and adaptive weighting based on effect size), with optional parameters selected through cross-model selection (Privé et al., 2022).
3. LDpred2 (Privé et al., 2020) [PGS004969, PGS004970, PGS004971, PGS004972, PGS004973, PGS004974 (Gunn et al., 2025)]: Bayesian shrinkage method that computes posterior mean SNP effect sizes by combining GWAS summary statistics with LD information from an external reference population.
4. LDpred2-auto (PGS002150 (Privé et al., 2022)]: PRS were built by combining GWAS summary statistics and UK Biobank-based LD information and letting the method automatically estimate the proportion of causal variants and strength of effects. They also improved accuracy by correcting for large-effect variants and winner’s curse, and by focusing on the top most informative variants (Privé et al., 2022).
5. PolyFun-pred (PGS002654 (Weissbrod et al., 2020)]: Used genome-wide, functionally informed fine-mapping to estimate causal SNP effects in order to improve prediction (across populations) by prioritizing variants that are more likely to be causal rather than relying on LD proxies (Weissbrod et al., 2022).
6. DBSLMM (Yang & Zhou, 2020) [PGS003037 (Ma et al., 2022)]: Deterministic Bayesian sparse linear mixed model, it first identifies large-effect SNPs via clumping and thresholding (C+T), then estimates both large and small SNP effects analytically to produce a PRS that does not require cross-validation (Ma et al., 2022).
7. PRS-CSx (Ruan et al., 2022) [PGS003869 (Shim et al., 2023), PGS004981 (Gunn et al., 2025)]: An extension of PRS-CS where multiple (more than one) external, population-specific, reference panels coupled with GWAS summary statistics are used to improve cross-population prediction (Ruan et al., 2022). PRS-CSx results in multiple population-specific PRSs, which are often combined into a single PRS as a weighted sum. The component, population-specific PRSs may also be used and may differ from corresponding PRS-CS PRSs that do not use multiple populations in the development process.

### Statistical Analysis

All reported values were weighted to adjust for complex survey design, sampling probability and non-response in the HCHS/SOL cohort. Descriptive statistics for baseline characteristics are reported by Mainland (Mexican, South American, Central American) and Caribbean (Cuban, Dominican, Puerto Rican) Hispanic/Latino background groups (**Table 2**). Frequencies and weighted percents of the sample, were calculated for categorical variables.

**Table 2.**
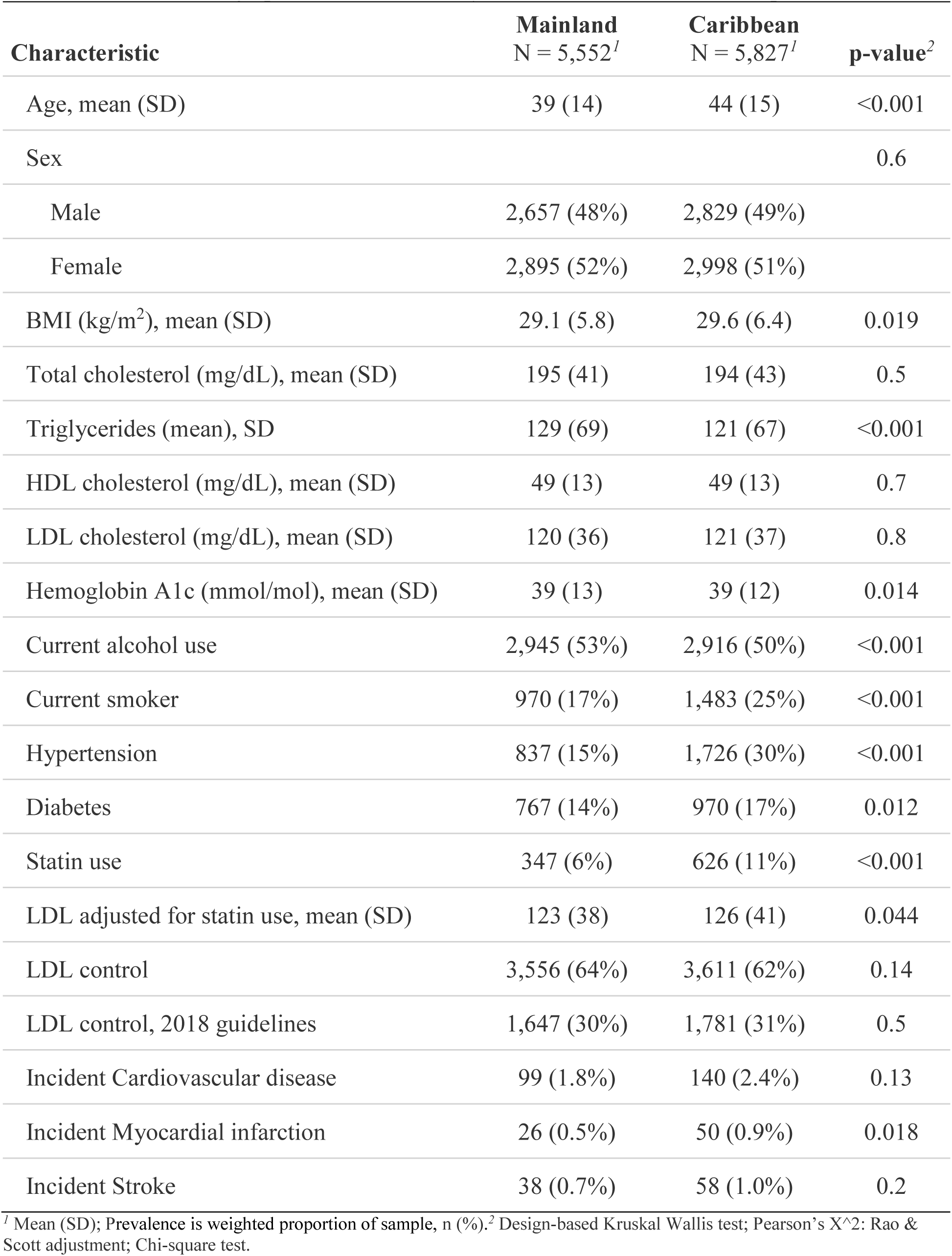
Baseline demographic characteristics by Mainland and Caribbean Hispanics/Latinos.

| Characteristic | Mainland<br>N = 5,552<sup>1</sup> | Caribbean<br>N = 5,827<sup>1</sup> | p-value<sup>2</sup> |
| --- | --- | --- | --- |
| Age, mean (SD) | 39 (14) | 44 (15) | <0.001 |
| Sex |  |  | 0.6 |
| Male | 2,657 (48%) | 2,829 (49%) |  |
| Female | 2,895 (52%) | 2,998 (51%) |  |
| BMI (kg/m<sup>2</sup>), mean (SD) | 29.1 (5.8) | 29.6 (6.4) | 0.019 |
| Total cholesterol (mg/dL), mean (SD) | 195 (41) | 194 (43) | 0.5 |
| Triglycerides (mean), SD | 129 (69) | 121 (67) | <0.001 |
| HDL cholesterol (mg/dL), mean (SD) | 49 (13) | 49 (13) | 0.7 |
| LDL cholesterol (mg/dL), mean (SD) | 120 (36) | 121 (37) | 0.8 |
| Hemoglobin A1c (mmol/mol), mean (SD) | 39 (13) | 39 (12) | 0.014 |
| Current alcohol use | 2,945 (53%) | 2,916 (50%) | <0.001 |
| Current smoker | 970 (17%) | 1,483 (25%) | <0.001 |
| Hypertension | 837 (15%) | 1,726 (30%) | <0.001 |
| Diabetes | 767 (14%) | 970 (17%) | 0.012 |
| Statin use | 347 (6%) | 626 (11%) | <0.001 |
| LDL adjusted for statin use, mean (SD) | 123 (38) | 126 (41) | 0.044 |
| LDL control | 3,556 (64%) | 3,611 (62%) | 0.14 |
| LDL control, 2018 guidelines | 1,647 (30%) | 1,781 (31%) | 0.5 |
| Incident Cardiovascular disease | 99 (1.8%) | 140 (2.4%) | 0.13 |
| Incident Myocardial infarction | 26 (0.5%) | 50 (0.9%) | 0.018 |
| Incident Stroke | 38 (0.7%) | 58 (1.0%) | 0.2 |

<sup>1</sup> Mean (SD); Prevalence is weighted proportion of sample, n (%).<sup>2</sup> Design-based Kruskal Wallis test; Pearson's X<sup>2</sup>: Rao & Scott adjustment; Chi-square test.

Means and standard deviations (SD) were calculated for continuous baseline characteristics. Kruskal Wallis or Pearson’s Chi-square using Rao & Scott adjustment were used to measure statistical differences between Mainland and Caribbean background groups.

Survey-weighted generalized linear regression with a Gaussian distribution was used to evaluate the association between the PRS and adjusted LDL levels at baseline; effect estimates (betas (β)) and 95% confidence intervals (CI) were calculated. Survey designs were created for the full sample, for the Mainland subset, the Caribbean subset, and three additional subsets for those with a majority of European, African, or Amerindian genetic ancestries; where the majority of a genetic ancestry was defined as those in the top tertile of each genetic ancestry group. Survey designs were applied using the svyglm function from the ‘survey’ R package (Lumley, 2004).

Each PRS was modeled as a continuous variable and analyzed separately in each survey design subset. All models were adjusted for age, sex and the first 5 principal components (PCs) to account fo genetic ancestry and population structure (Conomos et al., 2016). Incremental R^2^ were calculated for each PRS and each survey design subgroup as the difference between the base model (age, sex and 5 PCs) and the full model (PRS, age, sex, and 5 PCs) to determine the added variance explained by each PRS and 95% confidence intervals were estimated using bootstrap replicated weights (n=100). (**Supplemental Table 1**).

Survey-weighted logistic regression models (quasibinomial family) for the Mainland and Caribbean subgroups were used to derive odds ratios (OR) and 95% CIs for the association between each PRS and LDL control in those using statins and were adjusted for age, sex and the first 5 PCs. The association of PRS with LDL control was explored for modification by diabetes status (Ciuculete et al., 2017) with the inclusion of a cross-product term and assessment of the interaction p-values.

All analyses were conducted using R version 4.3.2.

## Results

There were 11,669 participants in the analytic sample; mean (SD) age was 42 (15) and 52% were female (**Supplemental Figure 1, Table 2**). Compared to participants with Mainland ancestry (Mexican, South American, and Central American), participants with Caribbean ancestry (Cuban, Dominican, and Puerto Rican) were older, had higher BMI, lower triglycerides levels, and were more likely to smoke, have hypertension, and diabetes. While baseline LDL levels were similar between groups, adjusted LDL levels were higher (P=0.04) in the Caribbean subset who were also more likely to be statin users (P<0.001) (**Table 2)**. The prevalence of high LDL (>130mg/dL), after adjustment for statin use, was higher for males than females (41% vs. 37%). The prevalence of high total cholesterol increased with age and varied by Hispanic/Latino background group and field center, with highest prevalence in those with Cuban background (48%) and those from the Miami field center (46%) (**Supplemental Table 2)**.

The PRSs exhibited approximately normal distributions (**Supplemental Figure 2**). When stratified by Caribbean and Mainland subgroups, there was substantial overlap of distributions of each PRS between subgroups and distributions remained largely normal, although some showed modest differences in central tendency and variability (**Supplemental Figure 3**). The distributions of multiple PRS were shifted slightly towards higher mean values for the Mainland subgroup (**Supplemental Figure 3**). This remained true when further stratified by Hispanic/Latino background groups (**Supplemental Table 3**); each PRS mean was higher for those within the Mainland subgroup than the Caribbean subgroup with few exceptions. For example, the mean PRS values of PGS003869 for Central American participants were on average lower than Cuban participants [Beta (SD): -0.01 (0.86) vs. 0.04 (1.07), respectively] and the mean PRS values for PGS004970 for Central Americans (-0.14 (0.98) were on average lower than Puerto Ricans 0.07 (0.97) and Cubans 0.34 (1.04) (**Supplemental Table 3**).

All PRSs were positively associated with adjusted LDL levels at baseline (**Figure 1, Supplemental Figure 4**) in all subgroups. The majority of PRS conferred an increase in LDL levels by about 5-15 mg/dL for each SD increase in continuous PRS when adjusted for age, sex, and the first 5 principal components; however, the associations differed slightly by PRSs and subgroups. For example, two PRSs (PGS004969 and PGS004975) were associated with smaller LDL increases per SD of each PRS (by 3-7 mg/dL), while PGS002654 was associated with larger LDL increases, ranging from 10-15 mg/dL per SD (**Figure 1, Supplemental Figure 4**).

**Figure 1.**
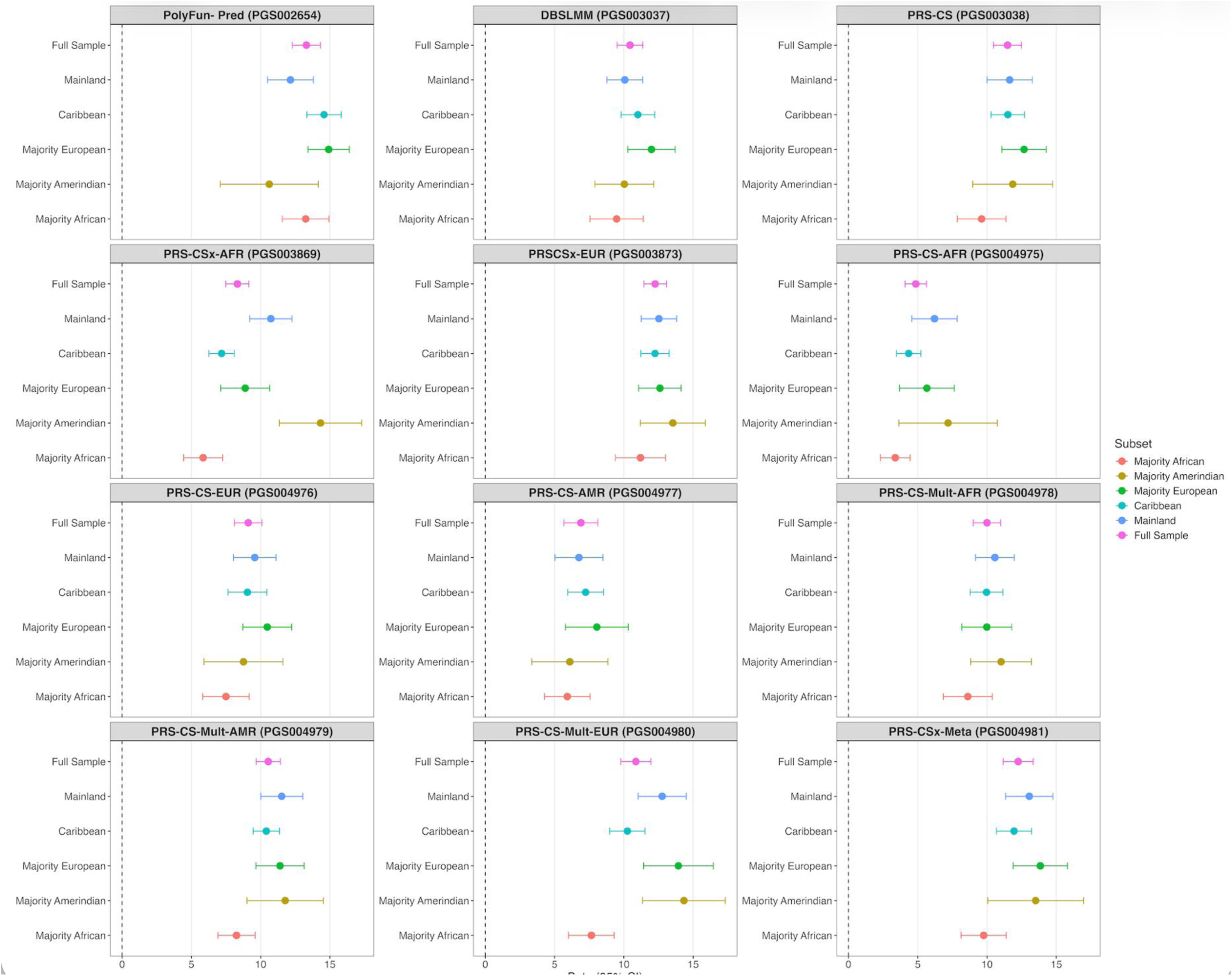
Forest Plot showing the associations for one standard deviation increases of each PRS with adjusted LDL levels across subgroups. Each PRS is shown separately across subgroups: Full sample (purple); Mainland (blue); Caribbean (teal); Majority European (green); Majoirty Amerindian (yellow); and Majority African (orange). Mainland subgroup consists of individuals of Mexican, Central American, and South American backgrounds. Caribbean subgroups consists of individuals of Cuban, Dominican, and Puerto Rican backgrounds. ‘Majority’ is defined as those in the top tertile of each genetic ancestry group. The association of one SD increase in PRS with LDL levels (mg/dL) adjusted by statin use, is reported as Betas (95% CI).

The incremental R^2^ for adding each PRS to the model with age, sex, and first 5 PCs was highest for PGS002654 in the full sample, Caribbean subgroup, those with a majority of African and Amerindian genetic ancestries (**Figure 2, Supplemental Figure 5**). The PRS that improved performance most for the Mainland subgroup and for those with a majority of European ancestry was PGS03873. For all subgroups, PGS004975 performed the worst (**Figure 2**). Sensitivity analysis assessed subgroup standardized PRS (Mainland and Caribbean standardized separately) for each PRS; however, PRS performance (R^2^) and the association of PRS with LDL levels (Betas) were not significantly changed. (**Supplemental Table 4)**.

**Figure 2.**
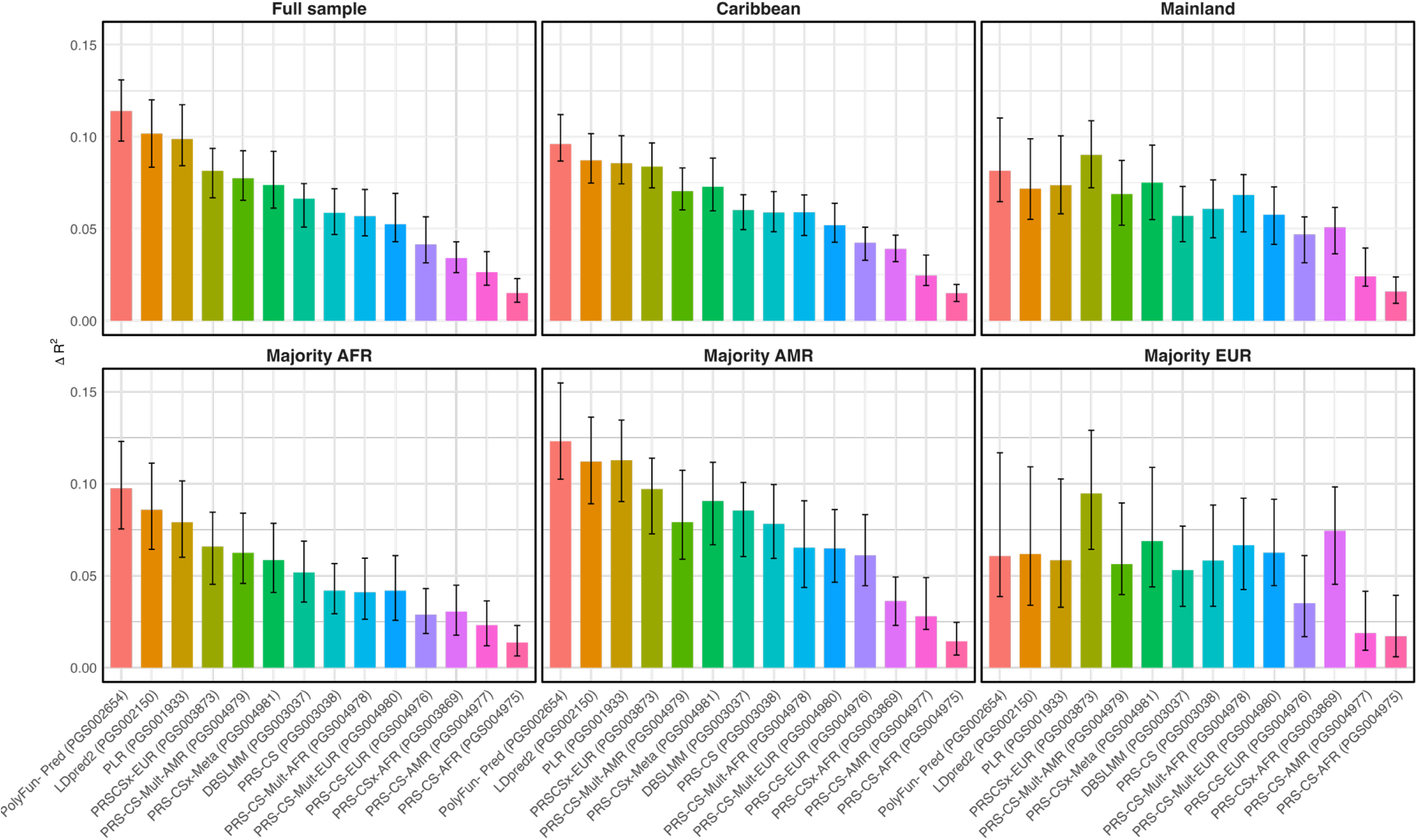
Incremental R^2^ comparing PRSs by subgroups. The change in R^2^ between a baseline model (age, sex, and 5 PCs) and a PRS model (baseline model + PRS) is reported for each PRS on the x-axis and delta R^2^ on the y-axis, plotted with bootstrapped confidence intervals. Each color corresponds to a different PRS.

We assessed each PRS for their association with LDL control among participants taking statins (n=1323) (**Figure 3, Supplemental Figure 6**). PGS002654 exhibited higher values of PRS to be associated with the lower odds of LDL control in both Caribbean and Mainland subgroups (**Figure 3**). In the Caribbean subgroup, continuous PGS002654 was associated with 33% (95% CI 0.55-0.82) lower odds of control for every one SD increase in PRS and 45% lower odds of control per SD increase in PRS for the Mainland subgroup (95% CI 0.38-0.79). PGS002654 provided nice delineation for odds of LDL control by increasing tertiles and quintiles of PRS for the Caribbean subgroup, while only the highest tertiles and highest quintiles showed significant associations with LDL control in the Mainland subgroup. (**Figure 3**). In the Mainland subgroup, PGS004978 showed a defined stepwise lower odds of control by increasing tertiles and quintiles of PRS (Figure 3). When assessing for differences in the association of PRS with LDL control by diabetes status, there was significant interaction between PGS004971 with diabetes (P=0.02) and PGS004977 with diabetes (P=0.02) only when using the updated definition of ‘control’ (LDL <100 or LDL <70 for those with diabetes (Grundy et al. 2019). For every one SD increase in PGS004971 the odds of control were lower for individuals without diabetes (OR 0.44[95% CI 0.27-0.74]) compared to those with diabetes (OR 0.88 [0.70-1.11]). Similarly, for every one SD increase in PGS004977, the odds of control were lower for individuals without diabetes (OR 0.52 [95% CI 0.33-0.82]) compared to those with diabetes (OR 0.93 [0.74-1.18]). **(Supplemental Table 5).**

**Figure 3.**
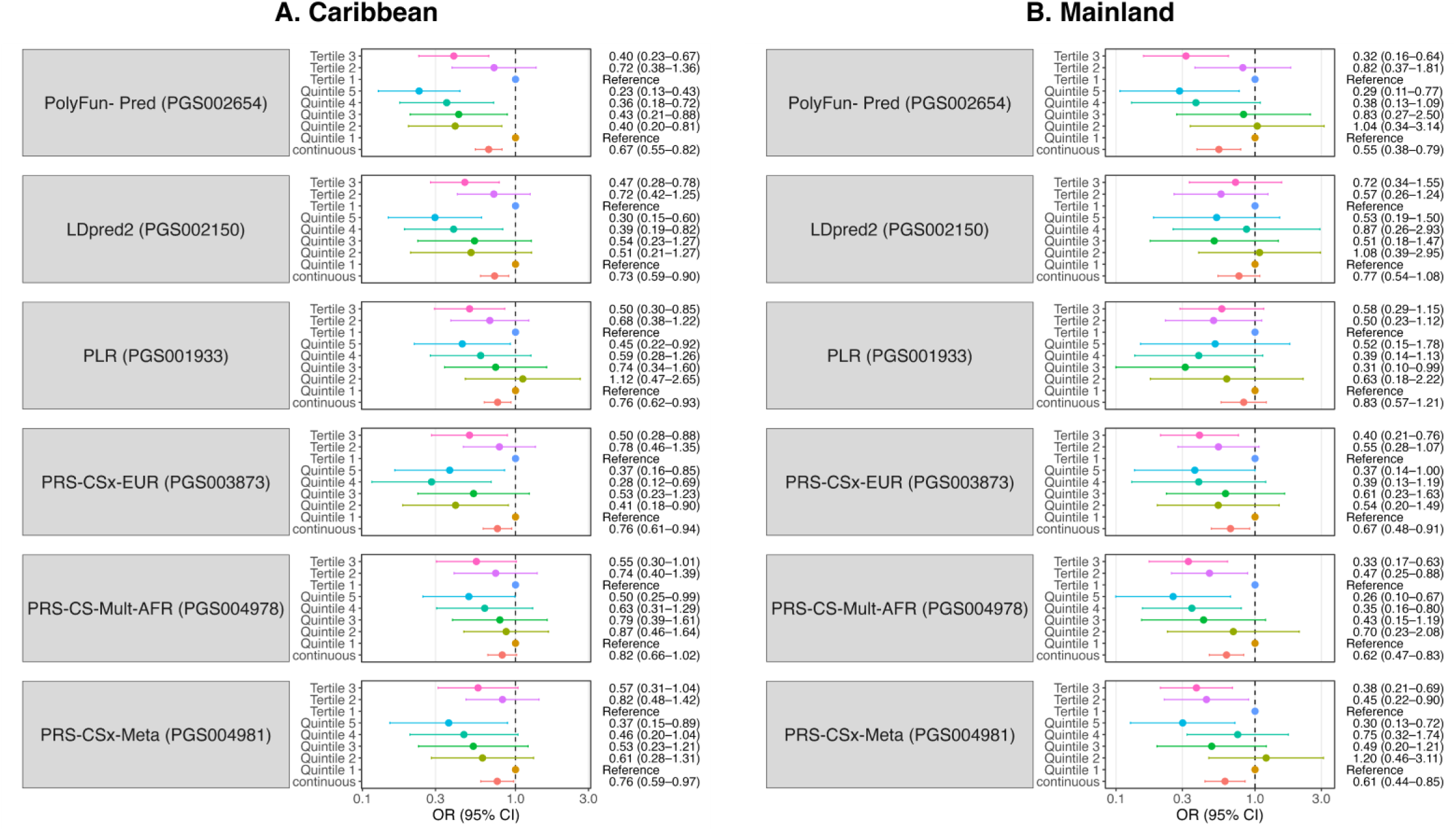
PRS associations with LDL control stratified by Caribbean and Mainland subgroups for those on statins. PRS are plotted by subgroups, A.) Caribbean and B.) Mainland. PRS are modeled continuously (red), by tertiles (pink and purple) and by quintiles (blues and greens). Odds ratios (95% Cis) are reported.

The association between the PRS and LDL control among statin users was stratified to analyze differences by the majority of European (n=285), African (n=381), and Amerindian (n=196) genetic ancestry for those with complete data on genetic ancestry, statin use, PRS, and model covariates (**Figure 4**). LDL control was defined as <130mg/dL or <100mg/dL for those with diabetes. PGS002654 remained associated with the lowest odds of control per SD increase in PRS only for those with a majority of European ancestry (OR 0.54, 95% CI 0.40-0.73) (**Figure 4**). While PGS002654 conferred 74% (95% CI 0.0.16-0.79) lower odds of control for those with a majority of Amerindian ancestry, other PRS conferred even lower odds of control (PGS003037, OR 0.30, 95% CI 0.14-0.61; PGS004980, OR 0.31, 95% CI 0.13-0.74; PGS004981, OR 0.32, 95% CI 0.16-0.64). No PRS were significantly associated with lower odds of control in those with a majority of African ancestry (**Figure 4**).

**Figure 4.**
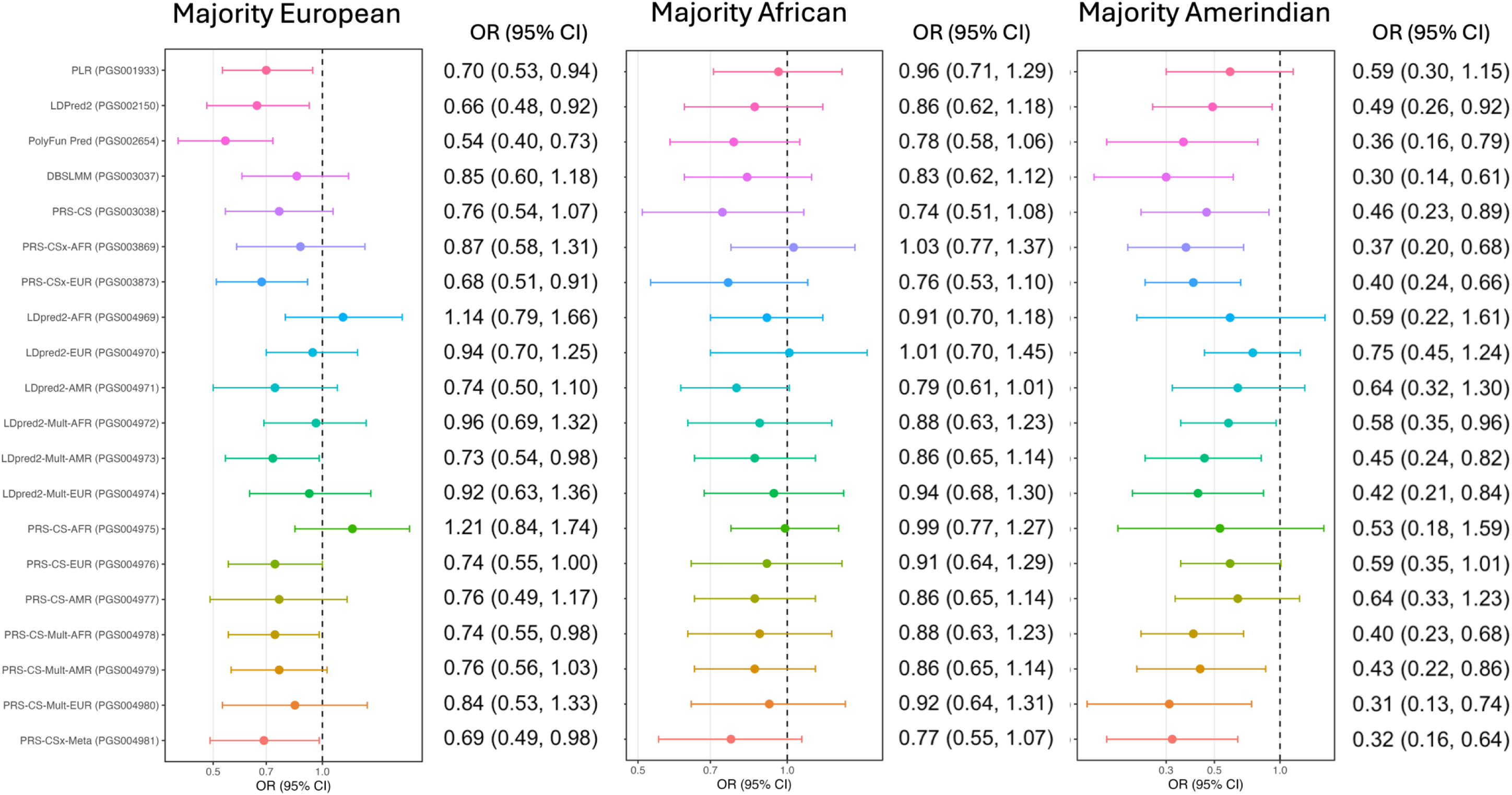
PRS associations with LDL control stratified by majority of genetic ancestry for those on statins. The association of each PRS was assessed separately for those with a majority of European (n = 285), African (n = 381), and Amerindian (n = 196). Models were adjusted for age, sex, and the first 5 PCs. LDL control was defined as <130mg/dL or <100mg/dL for those with diabetes. Each color corresponds to a different PRS.

## Discussion

We assessed multiple LDL-PRS for their association with LDL levels and with LDL control for those on statin therapy using Visit 1 data from the HCHS/SOL. Our findings show that higher scores of all PRSs were associated with higher LDL levels among all Hispanic/Latino subsets. Two PRS stood out, depending on genetic ancestry, as providing the most added value (incremental R^2^) to a base model adjusting for age, sex, and five PCs; specifically, PGS002654 in those with Caribbean background and PGS003873 in those with Mainland background performed best. The findings reinforce that differences in PRS performance exist across differing genetic backgrounds, certain methods may be superior for construction of PRS for use in a highly diverse and admixed population, and that caution should be used when choosing and applying PRS in Hispanic/Latino populations, as not all PRS are informative across subgroups for LDL levels and LDL control.

Trinder et al. found differences in the distributions of the PRS when stratifying by African, East Asian and European ancestries (Trinder et al., 2020). Although the magnitude of the difference in means was not discussed, the distribution of PRS values in participants with European genetic ancestry was shifted towards higher values compared to the sample of African participants (Trinder et al., 2020). Similarly, in this HCHS/SOL analysis, participants with Mainland-background (Mexican, Central American, or South American) on average exhibited the highest mean distribution, while the Caribbean-background groups (Puerto Rican, Cuban, and Dominican) showed lower mean distributions. Trinder et al. additionally showed that the magnitude of the association of the PRS with LDL levels was higher in individuals with European genetic ancestry (β[SE] 28.01[0.18]) compared to participants with African genetic ancestry (β[SE] 17.40[1.91]) (Trinder et al., 2020). While we had seen slight heterogeneity when stratifying the association of PRS with LDL in the HCHS/SOL cohort by subgroups, the more substantial differences in magnitude were seen across PRS.

According to improvements in incremental R^2^, PGS002654 performed best among the full sample and most subgroups (Caribbean, majority African and Amerindian genetic ancestries). It was also most informative for assessing odds of LDL control in all subgroups within HCHS/SOL, except for those with a majority of African genetic ancestry. PGS002654 was developed using PolyFun-pred and trained using European ancestry GWAS summary statistics from the UK Biobank (UKB). PolyFun-pred incorporates functional annotations to estimate posterior SNP effect sizes through Bayesian fine-mapping, prioritizing functional variants rather than those in LD with the causal SNPs (Weissbrod et al., 2022). By improving identification of likely causal variants, this approach can reduce the impact of differences in LD structure across populations and enhance the biological relevance of the resulting PRS (Schaid et al., 2018; Weissbrod et al., 2020). Previous utilization of PolyFun-pred illustrated improved prediction accuracy among Africans in the UKB over conventional LD-based approaches, consistent with our finding that PGS002654 exhibited the greatest predictive performance among the non-European subgroups of HCHS/SOL (Weissbrod et al., 2022). Functional annotations have been shown to improve performance of PRS for coronary heart disease in HCHS/SOL in the Caribbean subgroup, where utilizing AnnoPred (Hu et al., 2017) with and without traditional risk factors for CHD produced the highest concordance statistics compared to PRS developed using other advanced methods (Hutten et al., 2025).

PGS003873, which performed best among the Mainland subgroup and those with a majority of European genetic ancestry, was developed as part of PRS-CSx applied over African, Amerindian, East Asian, European, South Asian and Arab summary statistics, coupled with population-specific LD reference panels. (Shim et al., 2023). PRS-CSx results in population-specific PRSs corresponding to each of the population-specific summary statistics used for construction. Each population-specific PRS benefits from information borrowed across the multiple population-specific summary statistics; in that, the population-specific component PRSs extracted from PRS-CSx differ from population-specific PRSs developed using PRS-CS using the same population-specific summary statistics and LD reference panel. While PRS-CSx was purposely developed as an extension of PRS-CS in order to improve cross-population prediction by using external, population-specific, reference panels (Ruan et al., 2022; Shim et al., 2023), at time of publication, there were no available Hispanic/Latino GWAS summary statistics for LDL that do not include the HCHS/SOL cohort; therefore, we were unable to apply independent Amerindian-specific PRS-CSx in our analysis. We were able to use African-specific PRS-CSx (PGS003869), and while this method showed improvement over PRS-CS using African weights (PGS004975), both methods were among the least improved in the Caribbean and majority African genetic ancestry subgroups. Both PRSCSx-EUR (PGS003873) and PRS-CS-EUR (PGS004976) which used European GWAS summary statistics and weighting performed better than PRS-CSx-AFR (PGS003869) and PRS-CS-AFR (PGS004975) using African summary statistics and weighting in all subgroups. Surprisingly, the PGS003037 constructed using PRC-CS and European summary statistics from the UKB performed better than some PRS-CSx and PRS-CS models which used ancestry-specific and multi-ancestry summary statistics from the Million Veterans Program (MVP). This is in contrast to a study conducted in All of Us (AoU)

Research Program that found PRS-CSx improved performance over PRS-CS, regardless of Million Veterans Program (MVP) GWAS summary statistics used, including ancestry-specific GWASs and a fixed-effects meta-analysis across all ancestry groups (META GWAS) (Gunn et al., 2025). However, the diversity of the Million Veterans Program (MVP) Hispanic sample (e.g. Hispanic/Latino background group) is not reported and effect size for their Hispanic ancestry subset nearly matched the European subset of MVP (β = 10.5) suggesting their Hispanic sample originated from a population highly enriched for European ancestry (Graham et al., 2021).

For participants taking statins, we found that the best performing PRS in predicting LDL levels (PGS002654) was also highly associated with LDL control. PGS002654, developed using PolyFun-Pred, showed nice dose response in the association between PRS and LDL control, where higher tertiles and quintiles of PRS were associated with lower control in both the Caribbean subgroup and the Mainland subgroups. A study using data from the UK Biobank, of 33,787 adults using LDL lowering treatment, found a similar association between a 223-SNP PRS (Trinder et al., 2019) and odds of cholesterol control (Tapela et al., 2022). The odds of having uncontrolled hypercholesterolemia (defined as LDL >=3 mmol/L or 116mg/dL) were 2.78 times higher (95% CI 2.58, 3.00) for those in the top quintile of GRS (Tapela et al., 2022).

Although the UK Biobank is a fairly homogeneous sample, it does help confirm that LDL-PRS may be useful for determining individuals who may require medication adjustments to reach control. Others have also reported that higher PRS was associated with lower likelihood of LDL control but details about diversity of the sample and PRS analyzed were not available (Hanks et al., 2026). Individuals with higher PRS may require more intensive treatment to control LDL levels (Tapela et al., 2022). Support has been voiced for higher intensity statin treatment for those with severe hypercholesterolemia or for those in a “higher risk” subgroup but would need assessment within a group at high polygenic risk (Benziger & Groth, 2020; Bittencourt & Cesena, 2019). Others support using PRS earlier in life to identify those that could benefit from LDL lowering treatment to reduce athersclerotic cardiovascular disease (Hanks et al., 2026).

Our analysis had some limitations. Firstly, we could not tease out whether participants are using statins for indications other than high LDL; therefore, we are making certain assumptions when including people on statins for management of LDL control. Secondly, we did not have data related to statin dosage or levels of medication adherence which would also be a factor in having uncontrolled cholesterol levels. Third, we are not able to determine whether uncontrolled cholesterol levels translate to higher risk of cardiovascular (CVD) events in our sample of Hispanics/Latinos. Additional follow-up data is needed from the HCHS/SOL cohort to amass enough CVD events for this analysis to be possible. Fourth, we cannot rule out the possibility that some HCHS/SOL AMR-summary statistics are incorporated within the AFR- or EUR-specific PRS-CSx constructed using the GLGC data; however, the proportion would be small and unlikely to contribute to overfitting due to this joint training. Finally, this is a cross-sectional analysis using baseline data from the HCHS/SOL cohort. However, given that PRSs are our exposures of interest, temporality should not be an issue. We would also like to point out the strengths of this analysis. This is a unique study population with a well-characterized, large sample of diverse Hispanics/Latinos with genetic data. We were able to provide evidence for heterogeneity of associations between PRS and LDL levels and LDL control based on proportions of admixture among the Hispanics/Latinos living in the U.S., which has not been illustrated previously. Our analyses add to the PRS literature from a diverse Hispanic/Latino population that has been previously underrepresented in this area of research.

Our findings demonstrate that PRS for LDL vary substantially in performance across Hispanic/Latino populations, with the PRS which incorporated and prioritized functional variants (PolyFun-pred) providing the greatest predictive value for both LDL levels and LDL control among statin users. These results underscore the importance of testing PRS in diverse and admixed samples, as methods optimized in European or other populations may not translate effectively to more heterogeneous groups. While PRS show promise for identifying individuals at higher genetic risk who may benefit from earlier or more intensive lipid-lowering strategies, their clinical utility in diverse populations will depend on continued refinement using representative data and careful evaluation across subgroups. Future work incorporating longitudinal outcomes and treatment data with be essential to determine whether PRS can meaninguflly improve cardiovascular risk prediction and guide personalized care in Hispanic/Latino populations.

## Acknowledgements

CGH was supported by the National Heart, Lung, and Blood Institute (NHLBI) of the National Institutes of Health (NIH) under award numbers T32HL125294 (principal investigator[PI]: ML Daviglus) and F31HL154570 (PI: CGH), and by NIH/NHLBI award number T32HL007853 (PI: DJ Pinsky). The HCHS/SOL is a collaborative study supported by contracts from the NHLBI to the University of North Carolina (HHSN268201300001I/N01-HC-65233), the University of Miami (HHSN268201300004I/N01-HC-65234), the Albert Einstein College of Medicine (HHSN268201300002I/N01HC-65235), the University of Illinois at Chicago (HHSN268 201300003I/N01HC-65236), Northwestern University, and San Diego State University (HHSN268201300005I/N01-HC65237). The following institutes/centers/offices have contributed to the HCHS/SOL through a transfer of funds to the NHLBI: National Institute on Minority Health and Health Disparities, National Institute on Deafness and Other Communication Disorders, National Institute of Dental and Craniofacial Research, National Institute of Diabetes and Digestive and Kidney Diseases, National Institute of Neurological Disorders and Stroke, and the NIH Office of Dietary Supplements.

## Disclosures

The authors declare no competing interests.

## Data and code availability

HCHS/SOL data are available through application to the Database of Genotypes and Phenotypes (dbGaP: phs000810.v2.p2) or via a data use agreement with the HCHS/SOL Data Coordinating Center at the University of North Carolina at Chapel Hill (see collaborators’ website: https://sites.cscc.unc.edu/hchs/).

Code is available upon request.

## Supplemental information

See attached files.

## Web resources

PGS catalog, http://www.pgscatalog.org

## Notes

### Competing Interest Statement

The authors have declared no competing interest.

### Clinical Trial

N/A

### Author Declarations

Institutional Review Board (IRB) approval was obtained at each center's respective IRB, and all participants provided written informed consent in the language of their preference (English or Spanish).

